# Simulated identification of silent COVID-19 infections among children and estimated future infection rates with vaccination

**DOI:** 10.1101/2021.01.06.21249349

**Authors:** Seyed M. Moghadas, Meagan C. Fitzpatrick, Affan Shoukat, Kevin Zhang, Alison P. Galvani

**Author notes:** **Corresponding author:** Alison P. Galvani, Center for Infectious Disease Modeling and Analysis (CIDMA), Yale School of Public Health, New Haven, Connecticut, USA.

## Abstract

**Importance:** A significant proportion of COVID-19 transmission occurs silently during the pre-symptomatic and asymptomatic stages of infection. Children, while being important drivers of silent transmission, are not included in the current COVID-19 vaccination campaigns.

**Objective:** To investigate the benefits of identifying silent infections among children as a proxy for their vaccination.

**Design:** This study used an age-structured disease transmission model, parameterized with census data and estimates from published literature, to simulate the synergistic effect of interventions in reducing attack rates over the course of one year.

**Setting:** A synthetic population representative of the United States (US) demographics.

**Participants:** Six age groups of 0-4, 5-10, 11-18, 19-49, 50-64, 65+ years based on US census data.

**Interventions:** In addition to the isolation of symptomatic cases within 24 hours of symptom onset, vaccination of adults was implemented to reach a 40%-60% coverage over the course of one year with an efficacy of 95% against symptomatic and severe COVID-19.

**Main Outcomes and Measures:** The combinations of proportion and speed for detecting silent infections among children which would suppress future attack rates below 5%.

**Results:** In the base-case scenarios with an effective reproduction number *R*_e_ = 1.2, a targeted approach that identifies 11% and 14% of silent infections among children within 2 or 3 days post-infection, respectively, would bring attack rates under 5% with 40% vaccination coverage of adults. If silent infections among children remained undetected, achieving the same attack rates would require an unrealistically high vaccination coverage (at least 81%) of this age group, in addition to 40% vaccination coverage of adults. The effect of identifying silent infections was robust in sensitivity analyses with respect to vaccine efficacy against infection and reduced susceptibility of children to infection.

**Conclusions and Relevance:** In this simulation modeling study of a synthetic US population, in the absence of vaccine availability for children, a targeted approach to rapidly identify silent COVID-19 infections in this age group was estimated to significantly mitigate disease burden. Without measures to interrupt transmission chains from silent infections, vaccination of adults is unlikely to contain the outbreaks in the near term.

**Key Points:** *Question:* What is the effect of a targeted strategy for identification of silent COVID-19 infections among children in the absence of their vaccination?

*Findings:* In this simulation modeling study, it was found that identifying 10-20% of silent infections among children within three days post-infection would bring attack rates below 5% if only adults were vaccinated. If silent infections among children remained undetected, achieving the same attack rate would require an unrealistically high vaccination coverage (over 80%) of this age group, in addition to vaccination of adults.

*Meaning:* Rapid identification of silent infections among children can achieve comparable effects as would their vaccination.

## Introduction

The ongoing COVID-19 pandemic has caused significant global morbidity and mortality.^1^ Public health interventions, including social-distancing, testing and contact tracing, and isolation of cases, have substantially reduced the spread of SARS-CoV-2.^2–4^ However, enhanced viral transmissibility due to the emergence of novel variants ^5–8^ and the erosion of support for prolonged mitigation measures have raised concerns about perpetual waves of COVID-19 outbreaks.

Global efforts to ameliorate the impact of this deadly disease have galvanized the development of a number of vaccines which have received emergency use authorization from regulatory bodies in several countries,^9^ including Pfizer-BioNTech and Moderna in the United States (US).^10,11^ Most clinical trials have followed FDA guidelines,^12^ prioritizing the evaluation of vaccine safety and efficacy in adults, as this population group has borne the majority of reported infections, severe illnesses, and deaths.^13–15^ Given the lack of vaccine safety and efficacy data for children,^16^ vaccination campaigns have been targeted towards adults (18 years and older) and those at high risk of infection and severe outcomes. Thus, non-pharmaceutical interventions will still be required for mitigating disease transmission among children.

Given that children are more likely to develop asymptomatic infection compared to other age groups,^17–20^ they can be important drivers of silent transmission.^21^ We developed an age-stratified SARS-CoV-2 transmission model to investigate the impact of a targeted strategy for identifying silent infections among this age group when only adults are vaccinated. We then calculated the proportion and the speed of identification required to suppress future attack rates below 5% and, alternatively, the vaccination coverage among children which could achieve the same goal.

## Methods

### Ethics Statement

This study used publicly available data and parameter estimates from previously published studies and did not require Ethics review or approval.

### Model structure

We modelled the transmission of SARS–CoV-2 by developing an age-structured compartmental model taking into account the natural history of disease as well as self-isolation and vaccination dynamics (Appendix, Table A1). The population was stratified into six age groups: 0-4, 5-10, 11-18, 19-49, 50-64, 65+ years, parametrized from US census data.^22^ Model parameterization was based on age-specific data regarding asymptomatic rates of infection and relative transmissibilities during different stages of infection.^23,24^ Contact rates between and within age groups were heterogeneous and derived from empirical studies of social mixing.^25,26^ Newly infected individuals moved from the susceptible stage to the latent stage, and proceeded to a communicable silent infection stage (i.e., either asymptomatic or pre-symptomatic). A proportion of infected individuals remain asymptomatic until recovery,^17–20^ while others develop symptoms following the pre-symptomatic stage. The average duration of these epidemiological stages and other age-specific relevant parameters are derived from publicly available sources and published estimates (Appendix Table A2). For the base-case, susceptibility to infection was constant across ages, but as a sensitivity analysis, we reduced susceptibility by half for children under 10 years of age.^27–29^

In our model, all symptomatic cases were identified and isolated within 24 hours following symptom onset. For isolation of silent infections, we varied the proportion identified and the time from infection to identification in the range 2 to 5 days, reflecting observed delays in testing and contact tracing. Isolated individuals limited their daily contacts to the age-specific rates reported during COVID-19 lockdown^25,26^ until the end of their infectious period.

In vaccination scenarios, we distributed vaccines over time among individuals older than 18 years of age from the onset of simulations. Give vaccine prioritization of high-risk groups, we assumed that 80% of individuals 50 years and older and 22% of adults aged 18-49 years would be vaccinated, resulting in an overall vaccine coverage of 40% among adults within 1 year.^30^ We then extended our analysis for higher vaccination coverages of adults up to 60%. The vaccine efficacy against developing symptomatic or severe disease post-vaccination was 95%, based on the results of phase III clinical trials.^31,32^ We also assumed that vaccine efficacy against infection was 50% lower than the efficacy against disease, but also considered a scenario with the same efficacy of 95% as a sensitivity analysis (Appendix).

We calibrated the transmission rate to an effective reproduction number *R*_e_ = 1.2, accounting for the effect of current non-pharmaceutical interventions and 10% pre-existing immunity in the population.^33^ To capture the age distribution of pre-existing population immunity, the outbreak was simulated to the time prior to vaccination. The age-specific infection rates were then derived when the overall attack rate reached 10%,^34–36^ corresponding distributions of which were used as the starting population for the vaccination model. We assumed that the transmission rate was identical for pre-symptomatic and symptomatic cases, but reduced by 74% on average for asymptomatic cases based on recent estimates of asymptomatic COVID-19 infectivity,^24^ and that recovered individuals were not susceptible to reinfection. We then conducted model simulations independently for each intervention scenario and calculated the attack rate as the proportion of individuals infected within 1 year. For the scenario without vaccination, we considered identification of silent infections among all age groups. When vaccination of adults was implemented, identification of silent infections was targeted towards only children with delays of 2-5 days post-infection. In this targeted approach, the proportion and the speed of identification required to suppress future attack rates below 5% were determined. In the absence of pre-existing immunity and vaccination, most populations experienced an attack rate in the range of 1-5% during the first wave of the COVID-19 pandemic. Therefore, we assumed that an attack rate of under 5% would be a reasonable threshold to consider for our analysis in the presence of pre-existing immunity and vaccination while other non-pharmaceutical interventions are accounted for by the effective reproduction number. For each scenario of a time delay to identification, we calculated the vaccine coverage of children that would be required in addition to vaccination of adults, in order to achieve a similar attack rate if efforts to identify silent infections were completely halted.

### Statistical analysis

Simulations were seeded with an initial case in each age group in the latent stage in a population of 10,000 individuals for a time horizon of one year. In each scenario, cumulative infections were averaged over 1000 independent replications where disease specific parameters were sampled from their respective distributions (Appendix, Table 2). Credible intervals (CrI) at the 5% significance level were generated using the bias-corrected and accelerated bootstrap method (with 500 replications).

## Results

### Identification of silent infections in the population

In the absence of vaccination and with *R*_e_ = 1.2, an overall attack rate of 10.8% (95% CrI: 10.5% − 11.2%) would be expected when no silent infections in the population are detected (Figure 1). If silent infections are identified within 2 or 3 days post-infection, a rapid decline in the attack rate can be achieved with isolation of a relatively small (≤15%) percent of silent infections, with diminishing returns as identification rates rise above 20% (Figure 1). However, with a further delay in identification, a significantly larger proportion of silent infections need to be detected to have a similar impact in reducing the attack rate. For instance, with 10% of silent infections identified in the population and isolated within 2 days of infection, the attack rate can be reduced to 3.4% (95% CrI: 3.2% – 3.5%). To achieve the same mean attack rate with delays of 3, 4, and 5 days, detection rates of 13%, 42%, and 98% for silent infections would be required, respectively.

**Figure 1.**
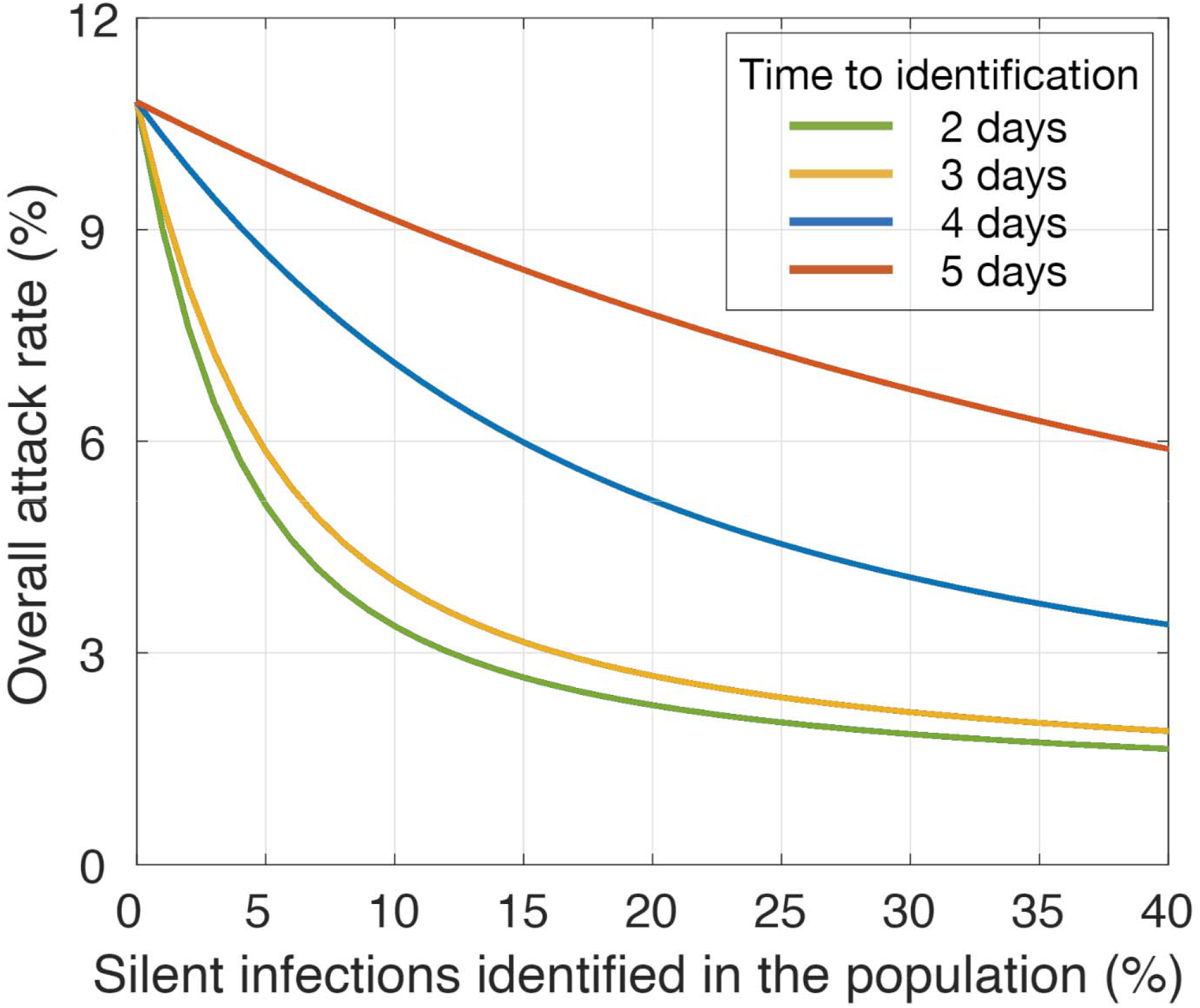
Estimated mean attack rates without vaccination and with identification of silen infections in the population. **Legend:** Colour curves represent attack rates when different proportions of silent infections are identified in the population, corresponding to different time delays post-infection.

### Targeted identification of silent infections among children

With vaccines distributed to only adults, attack rates among children and the overall population would be reduced to 12.5% (95% CrI: 11.9% – 13.2%) and 8.2% (95% CrI: 7.8% – 8.9%), respectively, without identification of silent infections (Figure 2). We simulated the effect of a targeted strategy for identification of silent infections only among children on reducing attack rates. Attack rates declined rapidly with increasing identification of silent infections within 2 or 3 days post-infection (Figure 2). For example, identification of at least 11% and 14% would suppress the overall attack rate below 5% (Figure 2B). With a delay of 4 and 5 days, significantly higher identification rates of 41% and 97%, respectively, are needed to bring attack rates under 5%. If silent infections among children remained undetected, an unrealistically high vaccination coverage (at least 81%) of this age group, in addition to 40% vaccination coverage of adults, must be achieved within 1 year to suppress attack rates below 5%. These results suggest that, even when vaccines do become available for children, rapid identification of their silent infections is still essential to mitigate disease burden in the population.

**Figure 2.**
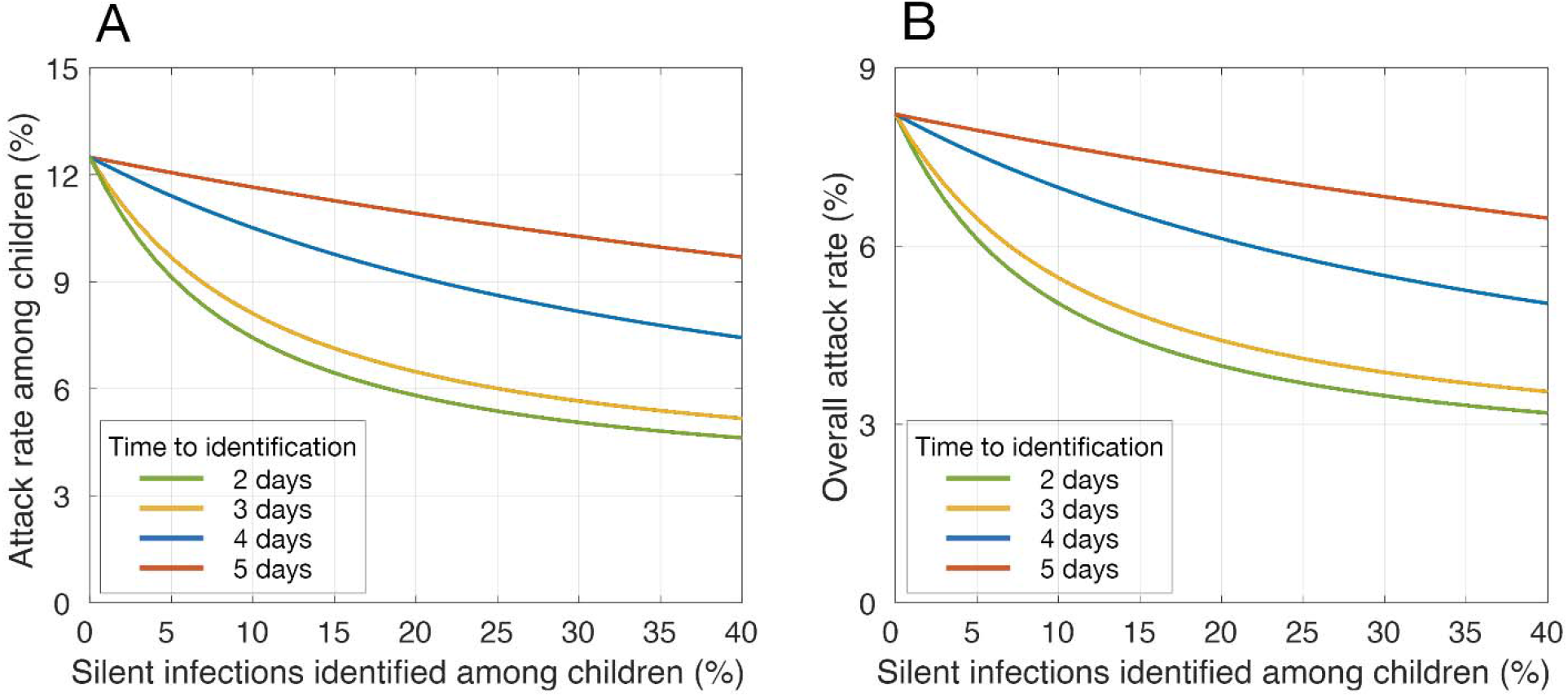
Estimated mean attack rates with vaccination of adults and identification of silent infections among children. **Legend:** Panels (A) and (B) represent attack rates among children (under 18 years of age) and the entire population, respectively. Colour curves represent attack rates when different proportions of silent infections are identified in children, corresponding to different time delays post-infection. Vaccination coverage of adults reached 40% within 1 year.

We further simulated the model to determine the effect of vaccine coverage on the minimum level of silent infections required to be identified among children in order to suppress the overall attack rate in the population below 5%. We found that when vaccination coverage of adults is expanded from 40% to 60%, the minimum identification levels dropped from 11% and 14% with delays of 2 and 3 days, respectively, to 5% and 6% (Figure 3). When the delay increased to 4 or 5 days, the minimum identification levels were 17% and 43% for a 60% vaccine coverage of adults over a one-year time-horizon, both of which were higher than those required for delays of 2 or 3 days with 40% coverage of adults.

**Figure 3.**
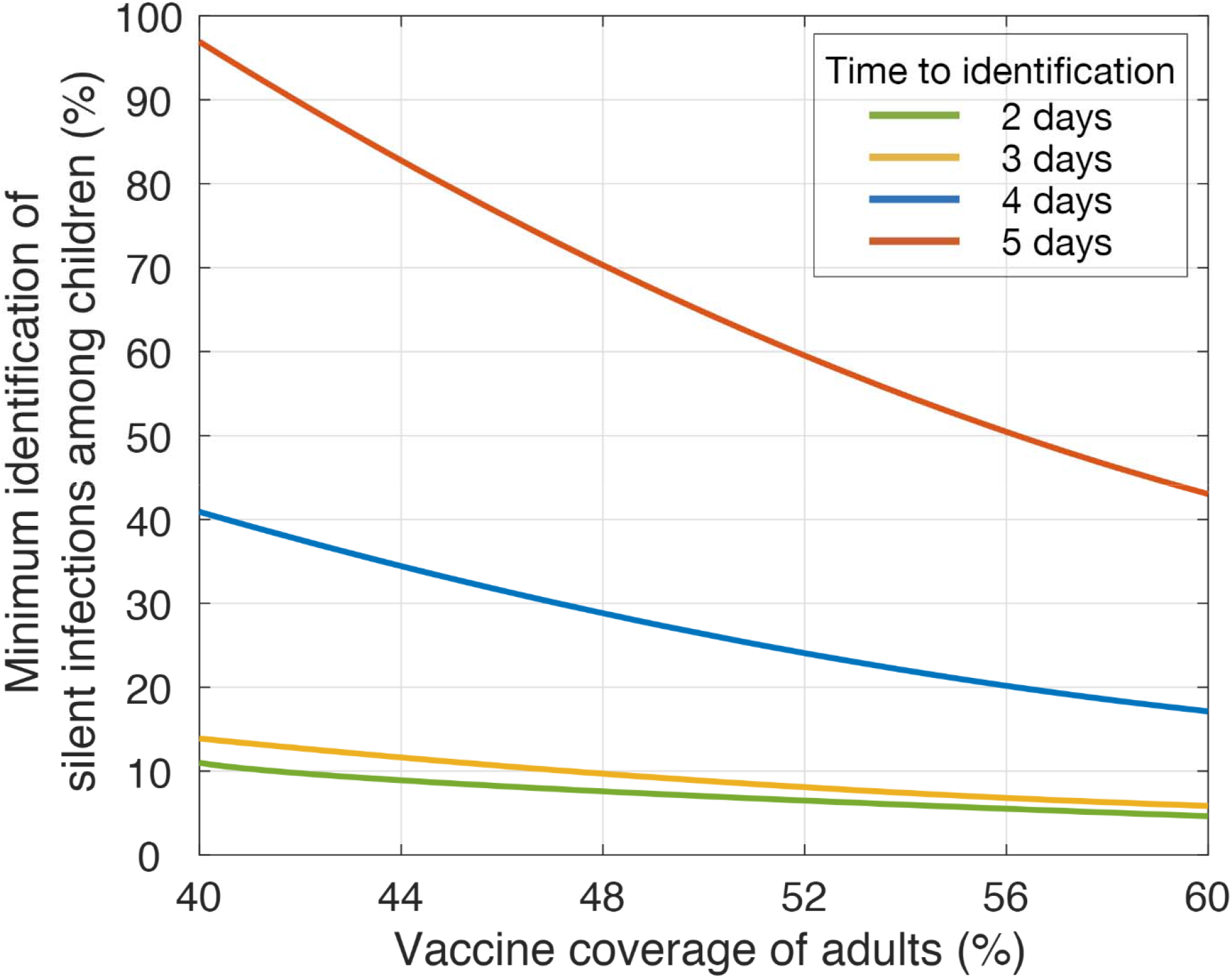
Required identification of silent infections among children to reduce attack rates below 5% with vaccination of adults. **Legend:** Colour curves represent the minimum identification level of silent infections among children required to suppress the overall attack rate below 5%, with different vaccination coverage of adults and time delays for identification post-infection.

### Sensitivity analyses

We evaluated whether reduced susceptibility to infection among children or higher vaccine efficacy against infection would affect the results. If susceptibility among children under 10 years old is reduced by half, then less contact-tracing is necessary to control COVID-19 with vaccination of adults (Appendix). For instance, 5%, 6%, 19%, or 47% identification of silent infections within 2, 3, 4, or 5 days following infection would suppress the overall attack rate below 5% (Appendix, Figure A2), or alternatively, vaccination coverage among children would need to reach 73% within 1 year. We observed qualitatively similar results when vaccine protection against infection was the same as efficacy against disease (Appendix). In addition, we conducted sensitivity analyses for a higher reproduction number of *R*_e_ = 1.5 and for a lower reproduction number of *R*_e_ = 0.9 to account for other factors such as seasonal effects that may influence disease transmissibility (Appendix). The results indicate that the identification of silent infections has a greater effect on reducing attack rates as the reproduction number increases.

## Discussion

A substantial proportion of COVID-19 cases are attributed to silent transmission from individuals in the pre-symptomatic and asymptomatic stages of infection.^37–40^ Children are particularly likely to have mild or asymptomatic infections,^18,41^ increasing the likelihood that they will serve as unidentified links between more severe cases. Although vaccines against COVID-19 now have emergency use authorization, these products have not yet been tested in children and it will be several months before children are widely vaccinated. In the absence of their vaccination, augmenting symptom-based screening with identification of silent infections is essential to control outbreaks.^23,42^ Our results suggest that the proportion of silent infections being identified among children is secondary to the speed of identification. For example, when *R*_e_ = 1.2, if the time from infection to identification was reduced from 4 days to 2 days post-infection without reduction of susceptibility for children under 10 years old, the same overall attack rate of 5% could be achieved with identifying over 3.7-fold (from 41% to 11%) lower proportion of silent infections. Accelerating identification from 5 days to 3 days corresponds to a 6.9-fold (from 97% to 14%) reduction in the proportion for detection of silent infections among children required to suppress the overall attack rate. Therefore, enhancing the capacity for rapid tracing of contacts of symptomatic individuals is critical to mitigating disease transmission.

The resurgence of COVID-19 cases prior to initiating vaccination in December 2020 overwhelmed the healthcare system in many jurisdictions, hampering the ability of public health to conduct effective contact tracing.^43–45^ Vaccination can alleviate the burden of COVID-19 outbreaks, and may allow for resource reallocation towards targeted contact tracing in settings where unvaccinated individuals congregate, such as schools and daycares. In a scenario where vaccines are only available for adults (with *R*_e_ = 1.2), our results show that if only 1 in 10 infected children were identified within 2 days post-infection, or 1 in 7 within 3 days post-infection (e.g., by contact tracing and routine testing), the overall attack rate could be reduced to below 5%. With recent advances in non-invasive testing modalities, such as saliva tests,^46^ routine testing in settings like schools could feasibly achieve this identification target.

### Limitations

Our results should be interpreted within the context of model limitations. First, we did not explicitly include the effects of non-pharmaceutical interventions, but instead calibrated the model to current estimates of the effective reproduction number which implicitly account for these effects.^33^ The relaxation of such measures would increase the need for vigilant contact-tracing among unvaccinated populations. Given COVID-19 awareness and public health recommendations, we assumed that all symptomatic cases self-isolate within 24 hours of symptom onset. Despite this high rate of rapid self-isolation, our sensitivity analyses confirm that rapid contact-tracing will still be an important dimension of control even if child susceptibility is half that of adults. With vaccination of adults, we evaluated the impact of identifying silent infections only among children. However, our results should not be interpreted as excluding adults for identification of silent infections. Our focus on targeting children is largely motivated by current deliberations regarding the reopening of schools and the potential for ensuing elevated spread of COVID-19 through asymptomatic infections in this population. Simultaneously expanding identification of silent infections among young adults currently not prioritized for vaccination would contribute to earlier control of outbreaks.

For the effect of vaccination, we parameterized the model with results of phase III clinical trials for vaccine efficacy.^31,32^ Given the uncertainty around distribution capacity and uptake of vaccines, we simulated the model with a vaccination rate to achieve 40%-60% vaccine coverage of adults within one year. If vaccines are distributed more rapidly or with higher uptake, it is possible that the rapid rise of population-level immunity could reduce the need for a targeted strategy to identify silent infections in children. However, given the current limitations in initial vaccine supplies and challenges with cold-chain distribution of mRNA vaccines,^47,48^ it is unlikely that vaccination will remove the need for non-pharmaceutical interventions in the near-term.

## Conclusions

In this simulation modeling study of COVID-19 transmission dynamics, identification of silent infections among children was shown to be an important strategy as vaccination campaigns continue to immunize adults. We found that early interruption of transmission chains is critical to outbreak control. Contact tracing at the time of symptom onset or testing, as opposed to at the time of testing results, could have a large impact on suppressing onward disease transmission by asymptomatic or pre-symptomatic infections, especially in the context of delays in turnaround time for COVID-19 test results.

## Data Availability

N/A

## Acknowledgement

Seyed M. Moghadas, Meagan C. Fitzpatrick, and Affan Shoukat contributed equally to this study.

## Author Contributions

Dr. Moghadas had full access to all the data in the study and takes responsibility for the integrity of the data and the accuracy of the data analysis.

*Concept and design:* Moghadas, Galvani.

*Acquisition, analysis, or interpretation of data:* All authors.

*Drafting of the manuscript:* All authors.

*Critical revision of the manuscript for important intellectual content:* Fitzpatrick, Moghadas.

*Statistical analysis:* Moghadas, Shoukat.

*Obtained funding:* Moghadas, Galvani.

*Administrative, technical, or material support:* Zhang.

*Supervision:* Moghadas, Galvani.

