## Supplemental File for "Simulated identification of silent COVID-19 infections among children and estimated future infection rates with vaccination"

#### The Model

We modelled the transmission of SARS–CoV-2 by extending an age-structured SEIR (Susceptible, Exposed, Infectious, Recovered) to include additional compartments of asymptomatic, pre-symptomatic, symptomatic, and isolation of infected individuals (Figure A1). We further included compartments to describe vaccination dynamics. The total population was divided into five age groups as specified in the main text. We omitted the demographic variables of births and deaths. With the variables described in Table A1, the model is expressed by the following system of equations:

$$
\begin{aligned}
S_a' &= -S_a\mathcal{J}_a - \xi_a S_a \\
V_a' &= \xi_a S_a - (1-\epsilon_a)V_a\mathcal{J}_a \\
E_a' &= (1-q_a)S_a\mathcal{J}_a - \sigma E_a \\
\mathcal{E}_a' &= (1-q_a)(1-\epsilon_a)V_a\mathcal{J}_a - \sigma\mathcal{E}_a \\
F_a' &= q_a S_a\mathcal{J}_a - \sigma F_a \\
\mathcal{F}_a &= q_a(1-\epsilon_a)V_a\mathcal{J}_a - \sigma\mathcal{F}_a \\
A_a' &= p_a\sigma E_a + \rho_a\sigma\mathcal{E}_a - (1-g_a)\eta A_a - g_a\delta A_a \\
P_a' &= (1-p_a)\sigma E_a + (1-\rho_a)\sigma\mathcal{E}_a - (1-g_a)\theta P_a - g_a\delta P_a \\
I_a' &= (1-g_a)\theta P_a - (1-f_a)\gamma I_a - f_a\tau I_a \\
G_a' &= p_a\sigma F_a + \rho_a\sigma\mathcal{F}_a - \eta G_a \\
H_a' &= (1-p_a)\sigma F_a + (1-\rho_a)\sigma\mathcal{F}_a - \left(\frac{\gamma\theta}{\gamma+\theta}\right)H_a \\
B_a' &= g_a\delta A_a - \left(\frac{\delta\eta}{\delta-\eta}\right)B_a \\
C_a' &= g_a\delta P_a - \left(\frac{\delta\theta\gamma}{\delta\theta+\gamma(\delta-\theta)}\right)C_a \\
Q_a' &= f_a\tau I_a - \left(\frac{\tau\gamma}{\tau-\gamma}\right)Q_a \\
R_a' &= (1-g_a)\eta A_a + (1-f_a)\gamma I_a + \eta G_a + \left(\frac{\gamma\theta}{\gamma+\theta}\right)H_a + \left(\frac{\delta\eta}{\delta-\eta}\right)B_a \\
&\quad + \left(\frac{\delta\theta\gamma}{\delta\theta+\gamma(\delta-\theta)}\right)C_a + \left(\frac{\tau\gamma}{\tau-\gamma}\right)Q_a
\end{aligned}
$$

with the force of infection given by

$$
\mathcal{J}_a = \beta\left(\sum_{j=1}^{6} M_{a,j}\frac{(P_j+\alpha A_j+I_j)}{N_j} + \sum_{j=1}^{6}\tilde{M}_{a,j}\frac{(C_j+\alpha B_j+Q_j+\alpha G_j+H_j)}{N_j}\right)
$$

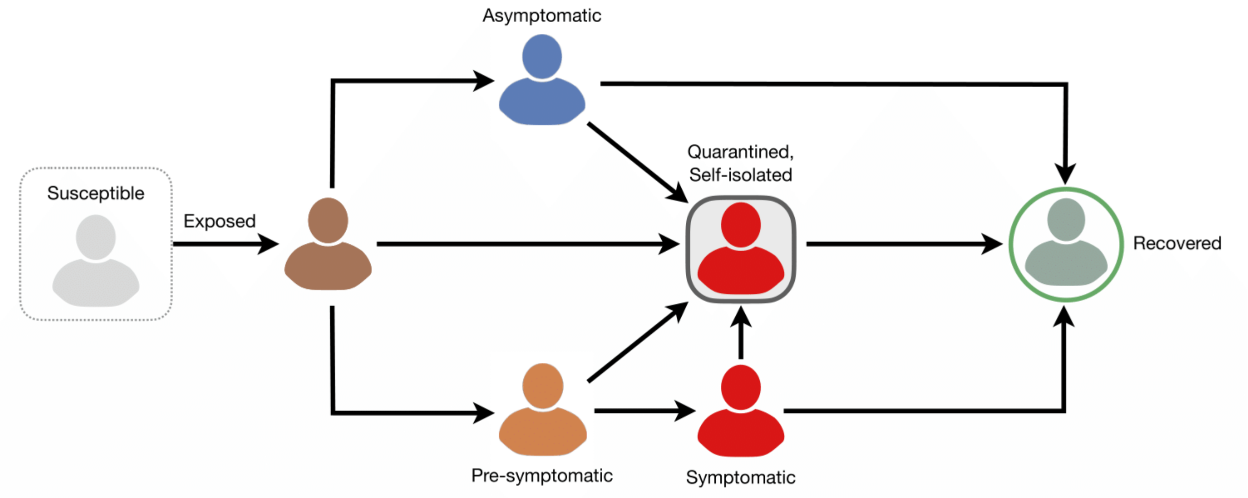


**Figure A1.** Schematic model diagram for disease transmission dynamics.

Table A1. Description of the model state variables.

| Variable | Description |
|---|---|
| $S_a$ | Susceptible in age group $a$ |
| $V_a$ | Vaccinated in age group $a$ |
| $E_a$ | Exposed in age group $a$ (without vaccination) |
| $\mathcal{E}_a$ | Exposed in age group $a$ (with vaccination) |
| $F_a$ | Identified within latent period in age group $a$ (without vaccination) |
| $\mathcal{F}_a$ | Identified within latent period in age group $a$ (with vaccination) |
| $A_a$ | Asymptomatic in age group $a$ |
| $P_a$ | Pre-symptomatic in age group $a$ |
| $I_a$ | Symptomatic in age group $a$ |
| $G_a$ | Asymptomatic isolated in age group $a$ directly from latency |
| $H_a$ | Pre-symptomatic isolated in age group $a$ directly from latency |
| $B_a$ | Asymptomatic isolated in age group $a$ |
| $C_a$ | Pre-symptomatic isolated in age group $a$ |
| $Q_a$ | Symptomatic isolated in age group $a$ |
| $R_a$ | Recovered in age group $a$ |
| $N_a$ | Population size of age group $a$ |

In this model, $\beta$ is the transmission parameter (calibrated to an effective reproduction number $R_e$. The reproduction number $R_e$ denotes the average number of secondary infections caused by an infected individual before recovering and becoming immune (or dying) in the presence of measures that aim to control disease spread. We calibrated the transmission parameter by calculating the spectral radius of the next-generation matrix [1]. A full description of all model parameters is given in Table A2. The population was stratified into six age groups: 0-4, 5-10, 11-18, 19-49, 50-64, 65+. Transmission between and within age groups was based on heterogeneous mixing with rates determined by age-specific contact matrices [2,3] for regular contacts $M$ and during isolation $\tilde{M}$:

| | 0-4 | 5-10 | 11-18 | 19-49 | 50-64 | 65+ |
|---|---|---|---|---|---|---|
| Age | | | | | | |

$$M = \begin{bmatrix} 2.34 & 2.35 & 1.88 & 4.31 & 1.14 & 0.55 \\ 0.41 & 0.41 & 8.83 & 4.26 & 0.88 & 0.43 \\ 0.46 & 0.46 & 10.02 & 4.83 & 0.99 & 0.49 \\ 0.51 & 0.52 & 2.01 & 8.63 & 1.96 & 0.68 \\ 0.27 & 0.27 & 1.23 & 5.48 & 3.07 & 1.21 \\ 0.16 & 0.17 & 0.87 & 3.26 & 1.75 & 1.96 \end{bmatrix} \begin{matrix} \text{0-4} \\ \text{5-10} \\ \text{11-18} \\ \text{19-49} \\ \text{50-64} \\ \text{65+} \end{matrix}$$

(Columns: 0-4, 5-10, 11-18, 19-49, 50-64, 65+; rows: Age)

and

(Columns: 0-4, 5-10, 11-18, 19-49, 50-64, 65+; rows: Age)

$$\tilde{M} = \begin{bmatrix} 0.64 & 0.65 & 0.53 & 1.21 & 0.32 & 0.15 \\ 0.11 & 0.12 & 2.3 & 1.21 & 0.25 & 0.12 \\ 0.12 & 0.13 & 2.8 & 1.35 & 0.28 & 0.14 \\ 0.14 & 0.15 & 0.56 & 2.41 & 0.55 & 0.19 \\ 0.07 & 0.08 & 0.34 & 1.53 & 0.86 & 0.34 \\ 0.05 & 0.05 & 0.24 & 0.91 & 0.49 & 0.55 \end{bmatrix} \begin{matrix} \text{0-4} \\ \text{5-10} \\ \text{11-18} \\ \text{19-49} \\ \text{50-64} \\ \text{65+} \end{matrix}$$

where in each matrix, the elements $\{m_{\mathrm{ij}} \mid i,j \in (1,\cdots,6)\}$ denote the average contact rates between age groups $i$ and $j$.

In our model, all newly infected individuals start in the latent stage for an average period of $1/\sigma$ days. After this period has elapsed, infected individuals move to a communicable silent infection stage (i.e., asymptomatic or pre-symptomatic). Unlike asymptomatic cases, those who enter pre-symptomatic stage will develop symptoms. We assumed that all symptomatic cases initiate self-isolation within 24 hours of their symptom onset. The average infectious periods in different stages of the disease and their associated distributions are summarized in Table A2. Recovery from infection was assumed to provide immunity against re-infection during the simulations.

To include vaccination dynamics, we considered age-dependent vaccination rates to achieve a 40% vaccine coverage in adults within 1 year, with a distribution of 80% for age groups 50+ and 22% for individuals aged 19-49. Vaccination was assumed to prevent infection with an efficacy that is 50% lower than its efficacy against symptomatic disease (and 95% in additional scenarios presented in as sensitivity analysis in this appendix). If infection occurred post-vaccination, we assumed the probability of developing symptomatic disease is reduced by a factor $\rho_{\mathrm{a}}$ corresponding to the vaccine efficacy of 95% [13].

For simulating the model, we used a non-standard numerical method to discretize the system and ran the simulations (in MATLAB©) with introducing one latent individual into each age group in the model. The time horizon of simulations was one year.

Table A2. Description of the model parameters and their associated values.

| Parameter | Description | Value | Source |
|---|---|---|---|
| $\beta$ | Transmission Parameter | Calibrated to $R_e$ | [4] |
| $\alpha$ | Relative transmissibility of asymptomatic infection | 0.26 | [5] |
| $1/\sigma$ | Mean latent period | 2.2 days | [6] |
| $q_a$ | % of infected individuals identified during latent period | 0% - 100% | Varied |
| $p_a$ | % of infected individuals that are asymptomatic | | |
| | Age Group 0 – 4 | 30% | [7] |
| | Age Group 5 – 10 | 30% | |
| | Age Group 11 – 18 | 37.3% | |
| | Age Group 19 – 49 | 32.8% | |
| | Age Group 50 – 64 | 32.8% | |
| | Age Group 65+ | 18.8% | |
| $g_a$ | % of infected individuals identified during asymptomatic and pre-symptomatic stages | 0% - 100% | Varied |
| $1/\eta$ | Mean infectious period of asymptomatic infection. | 5 days | [8.9] |
| $1/\delta$ | Time to identification of silent infections during asymptomatic and pre-symptomatic stages | 0.8 – 2.8 days | Assumed |
| $1/\theta$ | Mean duration of pre-symptomatic stage | 2.3 days | [10.11] |
| $f_a$ | % of symptomatic cases who self-isolate | 100% | Assumed |
| $1/\tau$ | Mean time to self-isolation post-symptom onset | 24 hours | Assumed |
| $1/\gamma$ | Mean infectious period post-symptom onset | 3.2 days | [8,9] |
| $\epsilon$ | Baseline vaccine efficacy in preventing disease | 95% | [11,12] |
| $\rho_a$ | % of vaccinated individuals who develop asymptomatic infection if infected post-vaccination | $p_a \leq \rho_a \leq 100\%$ | Calculated |
| $\xi_a$ | Vaccination rate, calculated to achieve coverage: | | |

| | |
|---|---|
| Age group: 0 – 18 (0% coverage) | 0 |
| Age group: 19 – 49 (22% coverage) | $7.935 \times 10^{-4}$/day |
| Age group: 50 – 64 (80% coverage) | $5.649 \times 10^{-4}$/day |
| Age group: 65+ (80% coverage) | $5.233 \times 10^{-4}$/day |

Distribution of the incubation period: $\mathrm{logNormal}(1.434, 0.661)$
Distribution of the pre-symptomatic period: $\mathrm{Gamma}(1.058, 2.174)$
Distribution of infectious period for asymptomatic infection: $\mathrm{Gamma}(5,1)$
Distribution of infectious period after the onset of symptoms: $\mathrm{Gamma}(2.768,1.1563)$

### Results with $R_e = 1.2$ and reduced susceptibility of children

Evidence is accumulating that young children may have a reduced susceptibility to SARS-CoV-2, with stronger immune responses that may prevent the development of symptomatic or severe disease [14,15]. We therefore simulated the model by considering a 50% reduction of susceptibility for children under 10 years of age. Qualitatively, the effect of identifying silent infections on the reduction of attack rates remains intact and the speed of identification is critical for outbreak control. Projected attack rates for the range of 2-5 days delay in identification of silent infections among children, when only adults are vaccinated, are presented in Figure A2. We also simulated the model to determine the effect of vaccine coverage on the minimum level of silent infections required to be identified among children in order to bring the overall attack rate in the population below 5% (Figure A3).

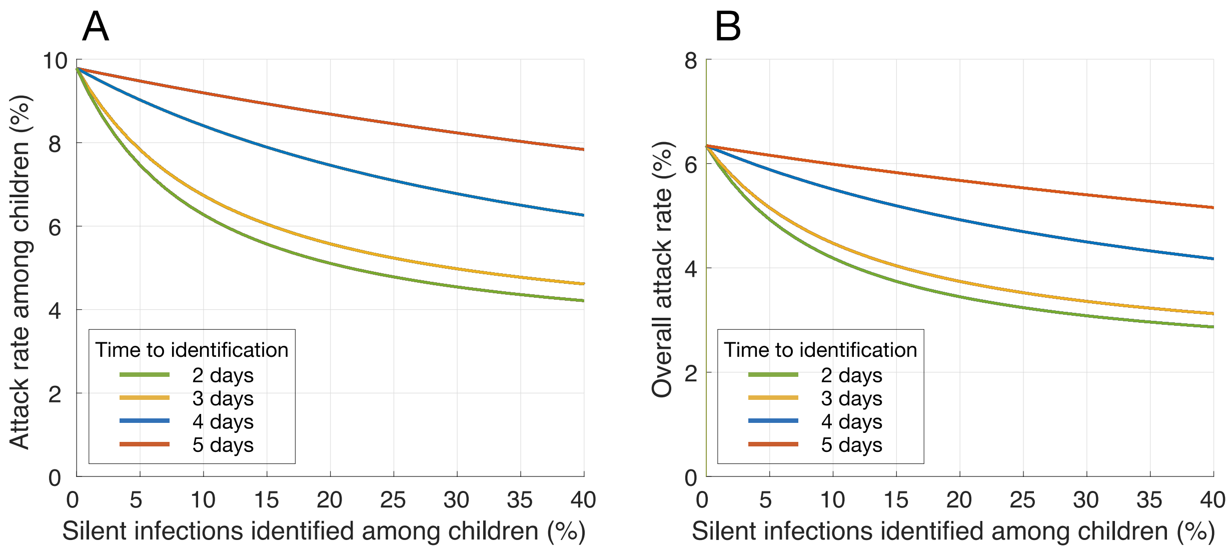


**Figure A2.** Estimated mean attack rate achieved with different rates of silent infections (i.e., asymptomatic and pre-symptomatic) identified and isolated among children, when only adults were vaccinated. Colour curves indicate the average time from infection to identification. Susceptibility of children under 10 years old was reduced by 50% compared to other age groups. Vaccine efficacy was assumed to be 95% against symptomatic disease, but 50% lower against infection. Vaccination coverage of adults reached 40% within 1 year.

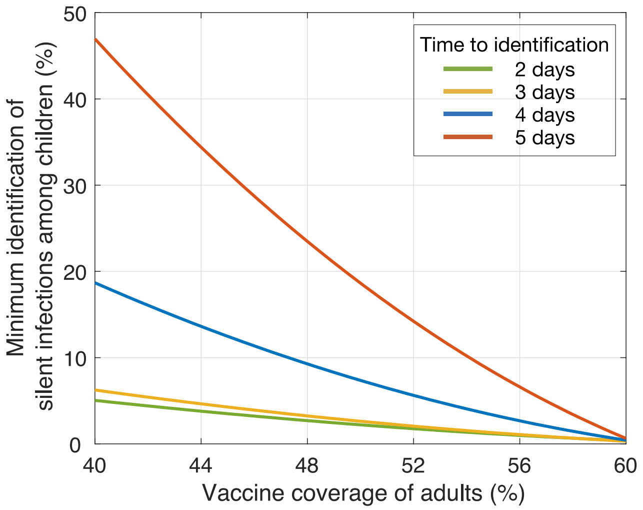


**Figure A3.** Minimum identification level of silent infections among children (y-axis) required to bring the overall attack rate in the population below 5% as a function of vaccine coverage of adults with different delays in identification post-infection.

#### Results with $R_e = 1.2$ and 95% vaccine efficacy against infection

In the absence of data on vaccine efficacy against infection, we further simulated the model with the same efficacy of 95% against symptomatic disease, while also considering 50% reduced susceptibility for children under 10 years old. The results presented in Figure A4 below illustrate a qualitative similar pattern to those presented in Figure A2 of the main text, indicating that the sharpest decline of attack rates occur with rapid identification of 0% - 15% silent infections among children within 2-3 days post-infection.

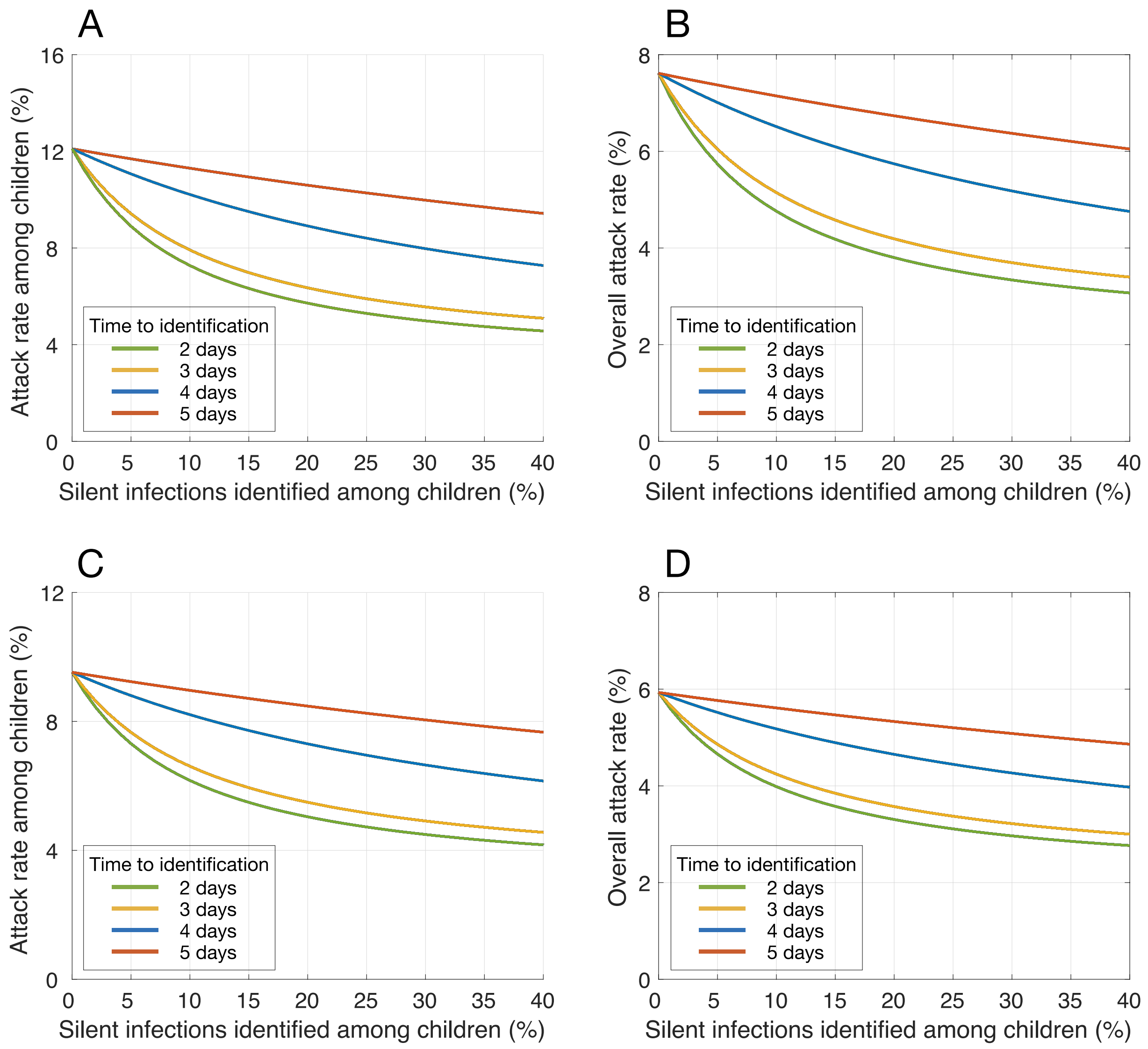


**Figure A4.** Estimated mean attack rate achieved with different rates of silent infections (i.e., asymptomatic and pre-symptomatic) identified and isolated among children, when only adults were vaccinated. Colour curves indicate the average time from infection to identification. Susceptibility of children under 10 years old was (A,B) the same as other age groups or (C,D) reduced by 50%. Vaccine efficacy was assumed to be 95% against both infection and symptomatic disease. Vaccination coverage of adults reached 40% within 1 year.

**Results with $R_e = 1.5$**

Depending on various factors (e.g., the characteristics of the disease, interventions, and other heterogeneities in the population), the reproduction number of diseases may change. As sensitivity analysis, we simulated the model when the reproduction number was increased to $R_e = 1.5$. Not surprisingly, attack rates were estimated to be higher and a greater proportion of silent infections in the population (without vaccination) and among children (with vaccination of adults) would need to be identified in order to suppress the overall attack rate below 5%. Figures A5-A7 show the results without vaccination, and when the vaccination coverage of adults is reached 40% over the course of 1-year. These simulations also consider reduced susceptibility of children under 10 years of age in scenarios with varying vaccine efficacy against infection (i.e., the same or 50% lower than the efficacy against symptomatic disease).

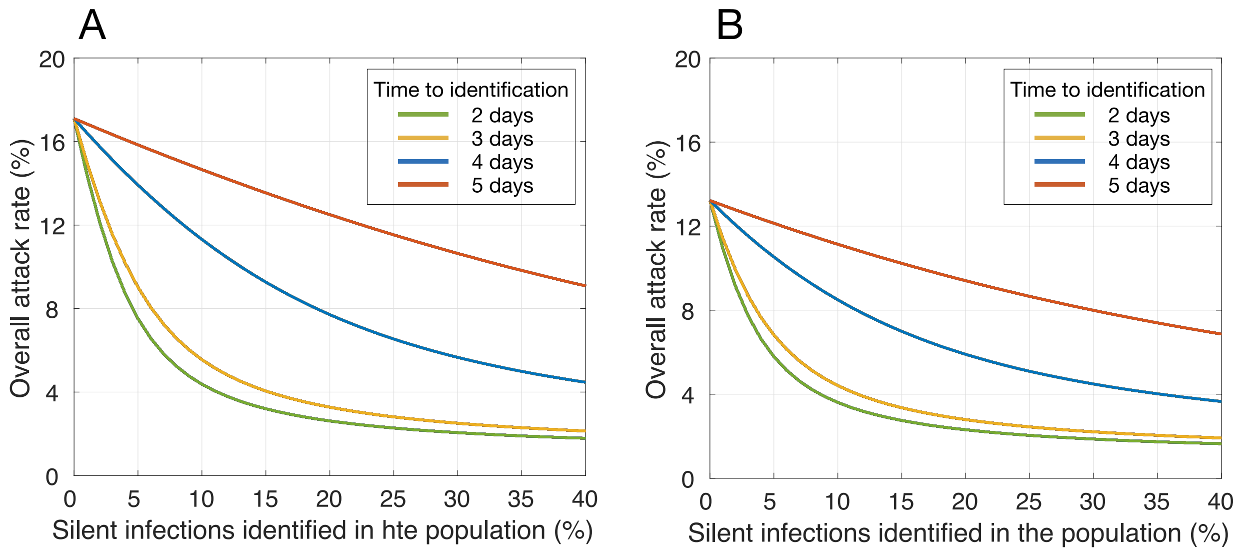


**Figure A5.** Estimated mean attack rate in the population achieved with different rates of silent infections (i.e., asymptomatic and pre-symptomatic) identified and isolated in the population without vaccination. Panel (A) and (B) correspond to full susceptibility and 50% reduced susceptibility of children under 10 years of age compared to other age groups. Colour curves indicate the average time from infection to identification.

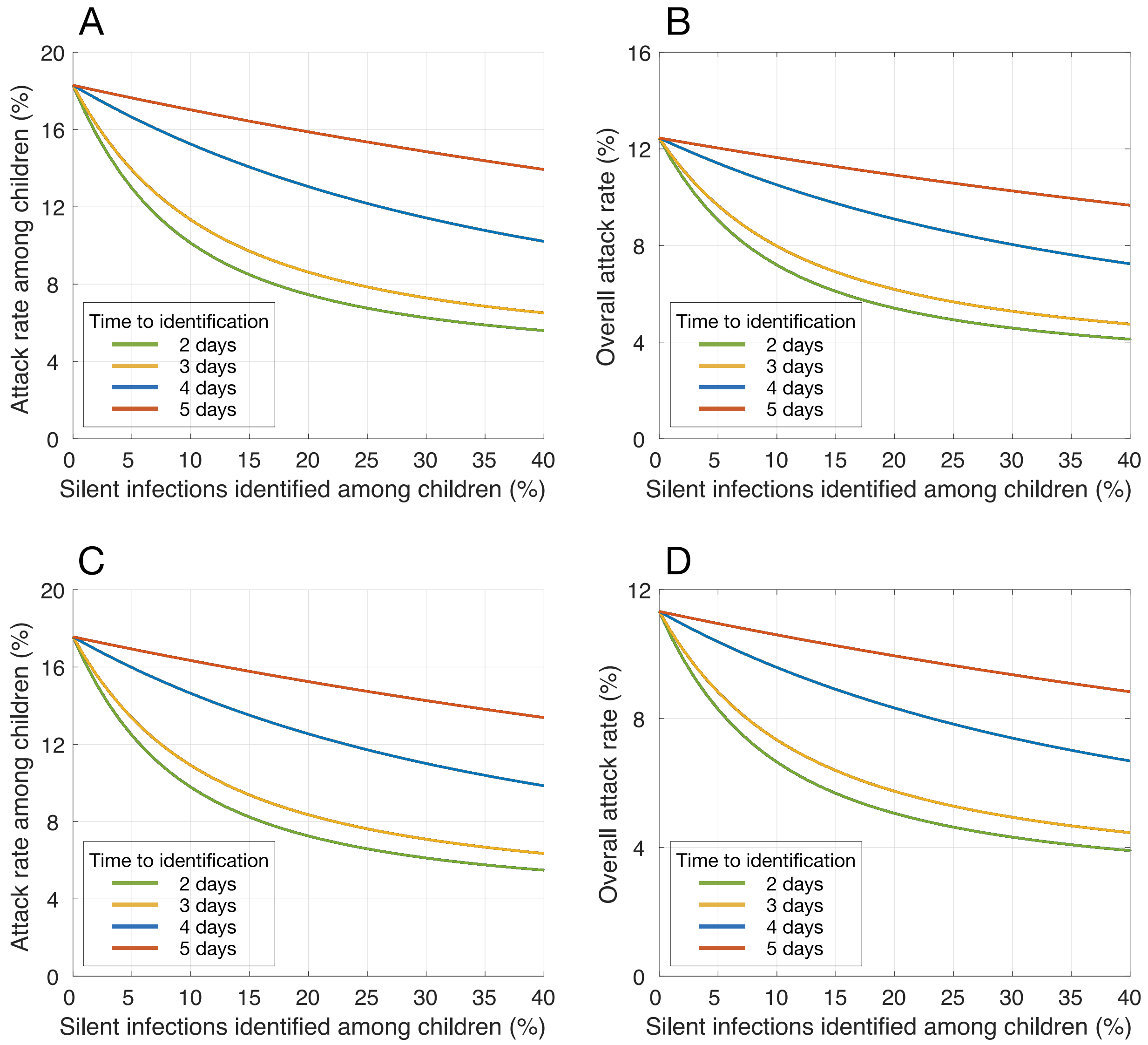


**Figure A6.** Estimated mean attack rate achieved with different rates of silent infections (i.e., asymptomatic and pre-symptomatic) identified and isolated among children, when only adults were vaccinated. Vaccine efficacy against infection is: (A,B) 50% lower than, or (C,D) the same as efficacy against symptomatic disease. Susceptibility of children under the age of 10 is the same other age groups. Colour curves indicate the average time from infection to identification. Vaccination coverage of adults reached 40% within 1 year.

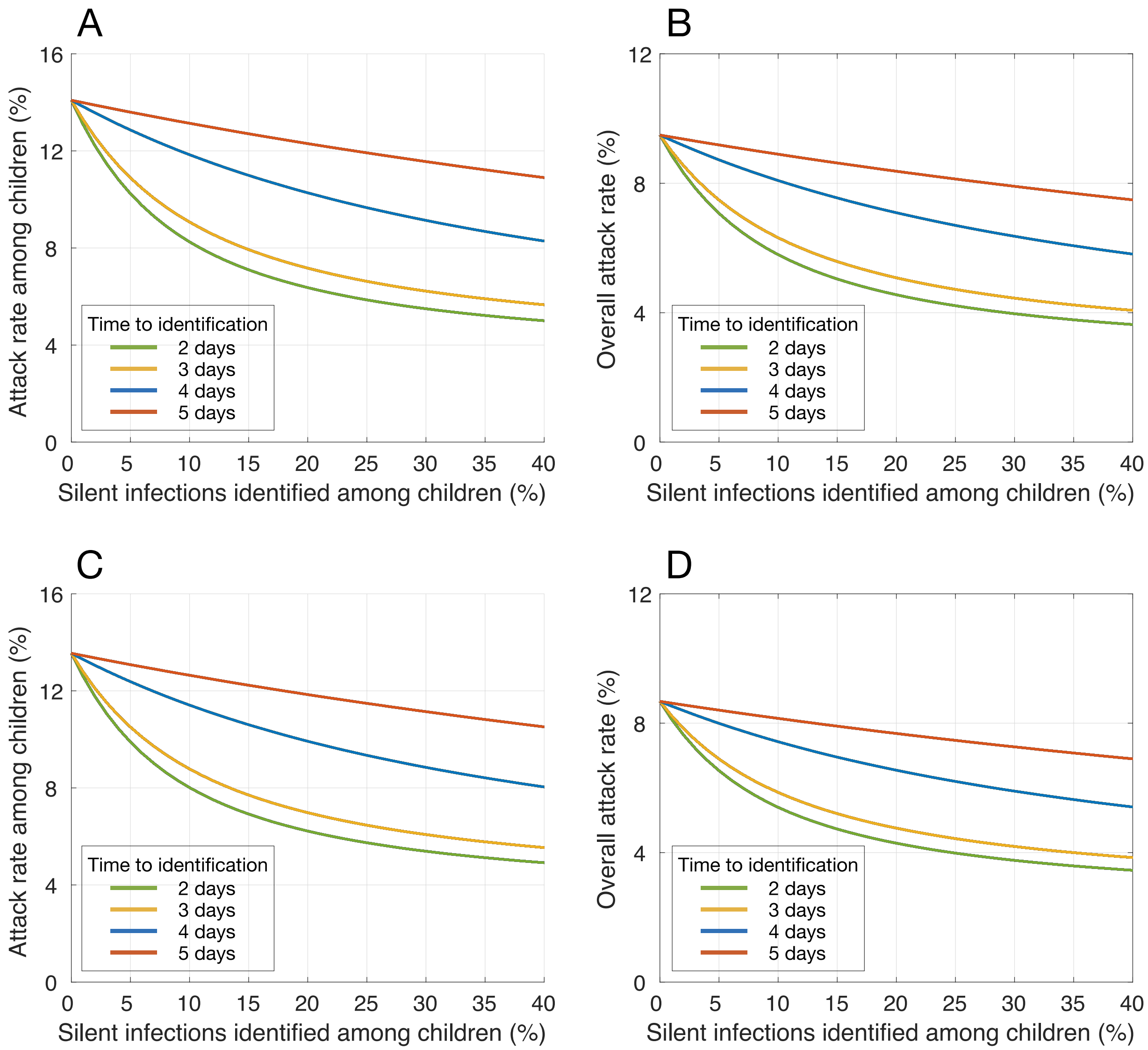


**Figure A7.** Estimated mean attack rate achieved with different rates of silent infections (i.e., asymptomatic and pre-symptomatic) identified and isolated among children, when only adults were vaccinated. Vaccine efficacy against infection is: (A,B) 50% lower than, or (C,D) the same as efficacy against symptomatic disease. Susceptibility of children under the age of 10 is 50% lower than other age groups. Colour curves indicate the average time from infection to identification. Vaccination coverage of adults reached 40% within 1 year.

#### Reduced reproduction number: $\boldsymbol{R_e = 0.9}$

When the reproduction was below one (simulated with $R_e = 0.9$), we found that with a 40% vaccine coverage, attack rates remained below 5% irrespective of the proportion of silent infection identified in the population or among children. However, as identification of silent infections increases with shorter delay post-infection, an earlier control of outbreak can be achieved.
